# “I was scared dating… who would take me with my status?”- Living with HIV in the UTT era in Johannesburg, South Africa

**DOI:** 10.1101/2022.07.29.22277160

**Authors:** Tembeka Sineke, Dorina Onoya, Idah Mokhele, Refiloe Cele, Shubhi Sharma, Smangele Sigasa, Mandisa Dukashe, Laila Hansrod, Robert Inglis, Rachel King, Jacob Bor

## Abstract

**BACKGROUND:** South Africa rolled out Universal Test-and-Treat (UTT) in 2016, extending treatment eligibility to all persons living with HIV (PLHIV). Through this study, we sought to understand the experience of people living with HIV in the UTT era in South Africa.

**METHODS:** In May 2021, we conducted in-depth interviews (IDI) (N = 27) with adult (≥ 18 years) PLHIV referred by HIV counsellors at three peri-urban primary healthcare clinics. We also conducted three focus group discussions (FGDs) (N = 27) with adult PLHIV recruited from clinics or from civil society organisations through snowball sampling. Follow-up interviews were conducted with 29 IDI and FGD participants, to gain a deeper understanding of their journey living with HIV. Participants were asked to reflect on their HIV diagnosis, what their HIV status meant to them in light of the UTT era and how, if at all, being HIV-positive affected their lives. Interviews and focus group discussions were audio-recorded, transcribed, translated to English, and analysed thematically.

**RESULTS:** The study included 4 men and 23 women recruited from clinics and 12 men and 16 women recruited from civil society (total N= 54). Participants reported that PLHIV could live a long life with antiretroviral therapy (ART) and that ART was widely accessible. However, they reported that HIV elicited feelings of guilt and shame as a sexually transmitted disease. Participants used the language of “blame” in discussing HIV transmission, citing their own reckless behaviour or blaming their partner for infecting them. Participants feared transmitting HIV to others and felt a responsibility to avoid transmission. To manage transmission anxieties, participants avoided sexual relationships, chose HIV-positive partners, and/or insisted on using condoms. Many participants feared – or had previously experienced – rejection by their partners due to their HIV status and reported hiding their medication, avoiding disclosure to their partners, or avoiding relationships altogether. Most participants also reported having low to no knowledge about treatment-as-prevention (TasP). Participants who were aware of TasP expressed less anxiety about transmitting HIV to others and greater confidence in having relationships.

**CONCLUSION:** Despite the normalization of HIV as a chronic disease, PLHIV still experience transmission anxiety and fears of rejection by their partners. Disseminating information on treatment-as-prevention could reduce the psychosocial burdens of living with HIV, encourage open communication with partners, and remove barriers to HIV testing and treatment adherence.

## INTRODUCTION

HIV/AIDS was imbued with social meaning from its earliest days, as people sought to understand this new, mysterious, and lethal condition. In 1981, United States public health officials infamously linked AIDS to four already-stigmatized “risk groups” – homosexuals, heroin users, haemophiliacs, and Haitians. As Susan Sontag wrote, AIDS diagnosis became a metaphor for indulgence, deviance, and transgression, a stigma or a mark of personal failure [1]. In sub-Saharan Africa (SSA), different metaphors emerged based on epidemiological and cultural contexts. AIDS was linked to sex work, witchcraft, divine punishment for sinful acts, and attributed to immorality [2-7]. The negative social meaning associated with HIV was rooted in real fears of early death, leaving loved ones behind, and contagion. Stigma, however, has been profoundly injurious to the mental health of people living with HIV (PLHIV) and has discouraged HIV testing, status disclosure, and treatment and prevention uptake [8-14].

Antiretroviral therapy (ART) has transformed HIV/AIDS into a clinically-manageable chronic disease with limited impact on life expectancy [15] and economic productivity roles [16, 17]. South Africa rolled out ART in the public sector in 2004, and approximately 5.1 million people were on therapy and virally suppressed in 2020[18]. Access to ART is widespread, with 96% of South Africans living within 10km of a health facility providing ART [19]. As of September 2016, all PLHIV are eligible for ART regardless of CD4+ lymphocyte count, under South Africa’s Universal Test-and-Treat (UTT) policy [20, 21].

Even though HIV is no longer viewed as a death sentence; HIV remains stigmatized [22-24]. A recent survey of young adults in rural South Africa found that 78% did not feel comfortable having a sexual relationship with someone who was HIV positive even if they were on ART [25]. These data are perhaps surprising given that people with HIV who are virally suppressed cannot transmit the virus sexually. Although the scientific basis for HIV treatment-as-prevention (TasP) was established in 2011, TasP has not historically been emphasized in HIV counselling in South Africa [26], resulting in low knowledge of TasP [25, 27]. Evidence from other contexts suggests that providing information on TasP, including the “Undetectable Equals Untransmittable (U=U)” message may alleviate stigma and improve self-image among PLHIV [28].

In this qualitative study, we sought to understand how PLHIV in Johannesburg, South Africa, experience and interpret their HIV status in the UTT era; how being HIV-positive affects their lives; and what role, if any, knowledge of TasP plays in their experience of living with HIV. Understanding the meaning ascribed to HIV and the challenges PLHIV face due to their status is critical to support psychosocial well-being and to motivate testing and ART adherence.

## METHODS AND MATERIALS

### Study design

This qualitative study was undertaken as formative research to guide the development and randomized evaluation of an intervention to integrate U=U/TasP into HIV counselling in South Africa (NIH R34 Bor/Onoya)[29, 30]. The larger project aims to determine whether informing patients about HIV treatment-as-prevention during HIV post-test and adherence counselling affects their knowledge and attitudes; stigma and wellbeing; as well as towards ART uptake and adherence. This formative research study consists of in-depth interviews and focus group discussions with PLHIV residing in and around Johannesburg, South Africa. Data was collected in May 2021.

### Data collection

Adults (_≥_ 18 years) living with HIV were recruited from peri-urban primary healthcare clinics in Johannesburg for participation in in-depth interviews. Potential participants were identified and referred by lay HIV counsellors. Trained study staff screened potential participants who were well enough to provide written informed consent in the participant’s preferred language. In addition, participants consented to have the findings written and published in scientific papers as part of the research work. Interviews lasted approximately 45 minutes and were conducted in a private space within the clinic. In-depth interviews covered the following domains: 1) perceptions of and experiences with HIV diagnosis (including thoughts and feelings after diagnosis, experiences with disclosure, and impact on relationships with family, friends and sexual partners), 2) In-depth understanding of challenges PLHIV experienced related to their HIV status and 3) perceptions about TasP. Interviews were conducted in English, Sotho and Zulu.

We also conducted three focus group discussions (FGDs). Participants for two of the FGDs were recruited through snowball sampling, starting with ART-experienced members of a civil society organisation that advocates for U=U/TasP in Johannesburg, South Africa. These FGDs were stratified by gender to promote open dialogue among participants around topics involving sexual relationships. A third FGD consisted of younger (**Table 1**.) and recently diagnosed participants recruited from clinics. The FGDs were facilitated by trained study staff and lasted for approximately two hours, covering similar topics to in-depth interviews including, experiences with the HIV diagnosis, experiences with HIV treatment, perceptions about treatment-as-prevention, and questions around communication methods for U=U/TasP.

**Table 1.** Characteristics of patients included in the analysis.

|  | Key informant interviews | Focus Group Discussions round 1 | Focus Group Discussions round 2 |
| --- | --- | --- | --- |
|  | N=27 (col%) | N=21 (col%) | N=6 (col%) |
| **Age at enrollment** |  |  |  |
| 18 - 30 | 24 (88.9) | N/A | 6 (100.0) |
| 30+ | 3 (11.1) | 21 (100.0) | N/A |
| **Gender** |  |  |  |
| Females | 23 (85.1) | 9 (42.9) | 6 (100.0) |
| Males | 4 (14.9) | 12 (57.1) | N/A |
| **Recruitment source** |  |  |  |
| Civil society organisation | N/A | 21 (100.0) | 6 (100.0) |
| Primary health facility | 27 (100.0) | N/A | N/A |

### Data analysis

Interviews and focus group discussions were audio-recorded, transcribed verbatim, translated to English, and analysed thematically. Data were analysed according to the following themes:

1. Meaning ascribed to HIV and PLHIV self-image in the UTT era
2. Challenges PLHIV experience related to their HIV status, particularly regarding relationships and disclosure and regarding treatment adherence
3. Understanding of U=U/TasP and its role in self-image and relationships

Major trends and cross-cutting themes were identified and then refined over several meetings. To maintain confidentiality and anonymity, all identifiers were removed from the final analytic data. This study was approved by the Human Research Ethics Committee (Medical) of the University of the Witwatersrand (M200529 MED20-05-019) and Boston University Medical Campus Institutional Review Board (H-40891).

## RESULTS

### 1. Meaning ascribed to HIV and PLHIV self-image in the UTT era

#### 1. Increased normalization of HIV

HIV was seen as “normalized” by most participants. And majority of the participants knew many people who were living with HIV. Treatment was ubiquitous, and HIV was no longer viewed as a death sentence. Many participants had friends and family members on ART, living healthy, normal lives:

> *“I can’t stop taking my medication because I am HIV positive, a lot of people are living normal lives, I’m not the only person and I’m not the first one*.*” –****Female***,***- 18-25yrs, key informant interview***

Participants did not view HIV as a death sentence with treatment widely available. Direct experience of family and friends having success with ART gave people confidence that they too could live a normal life with HIV:

> *“I have a younger sibling who was born with the virus from my parent, my sibling has been taking the treatment ever since. That is what gave me some motivation that if they could live for so long and be healthy then that means I can also do the same*.*” –* ***Female, 18-25yrs, key informant interview***

Despite the normalisation of HIV, participants reported feelings of guilt and shame related to their HIV diagnosis. In some cases, these feelings led to challenges in coping and accepting their HIV status and to delays in ART initiation:

> *“To be honest I was never okay, I even started drinking a lot and I didn’t take the treatment from September, October, November I think I only started taking the treatment in February this year*.*” –****Female, 18-29yrs, key informant interview***
>
> *“Umh like really, truly speaking; at first I was really disappointed as I am not that type who is busy dating all the time. I have one partner but then I learned to accept because I have kids, I had to tell myself that I have to live for their sake”*.*-* ***Female, 26-30yrs, key informant interview***
>
> *“I didn’t know how to feel. It was two different stories: I was pregnant and I was HIV+. For me, I felt disappointed, but then, I was just thankful I was still alive*.*”-* ***Female, 26-30yrs, key informant interview***

#### 1.2. Persistent HIV stigma is linked to sexual transmission

Despite the waning association of HIV with death, HIV still remained highly stigmatised as is often associated as a sexually transmitted disease. In addition to self-judgment, participants worried about the judgments of others. Being labelled as HIV-positive brings the most intimate aspects of one’s life into the public eye as HIV status disclosure involves the loss of privacy and control over information about one’s own private life and health. This internalized stigma negatively impacted acceptance of the diagnosis, with participants citing feelings of guilt and shame for contracting the virus. Some participants viewed their HIV diagnosis as an indicator of moral transgression. Participants described HIV transmission using the language of blame. Some blamed themselves for acting irresponsibly, while others blamed their partners, using language suggesting that HIV transmission indicated a breach of trust. One participant mourned her loss of virginity because of the resulting HIV infection, voices anger and frustration that she was not more “disciplined”:

> *“I* started *treatment in 2017 because I was in denial, was hiding it even at home, this thing and was scared to talk about this situation. Ok to think about it, the fact that I break my virginity when I was 21 isn’t. It means I had to discipline myself, thinking that you know what, unfortunately at the time I was breaking my virginity I got pregnant and I got HIV and it was so sad to me that I disciplined myself for such a long time I was well disciplined for this. I had -anger, I didn’t take treatment” –* ***Female, FGD, civil society group***

Another respondent blamed herself for what she felt was reckless behaviour and felt a strong motivation to avoid onward transmission:

> *“I took a risk, after taking that risk with my partner I saw him presenting HIV symptoms I knew it there and then that he is HIV positive because he had Shingles. So I don’t want to infect someone else” –****Female, 18-25yrs, key informant interview***

Participants described experiencing feelings of indignation at the carelessness of partners who did not try harder to protect them from HIV. They consequently felt a strong sense of responsibility to be careful and avoid transmitting HIV to others. Not having someone else to blame led to feelings of shame and forced people to reckon with their own role in transmission. Regardless of who was responsible in a given instance, a key theme was the use of language connoting culpability and blame.

> *“most embarrassing thing is not knowing who gave it to me*… *if I had someone to blame*,… *“but now I’m taking accountability*… *I was careless and don’t have anyone to blame” -* ***Female, 26-30yrs, key informant interview***

Transmission was interpreted as a breach of trust within a relationship.

> *“I didn’t believe it at first because I only had one partner. I trusted that person and I didn’t expect him to infect me with the virus. I was very confused because we have been together for years It took me a while to accept it. But as time went on, I ended up accepting” –****Female, 18-25yrs, key informant interview***

### 2. Challenges PLHIV experienced related to their HIV status regarding relationships and disclosure

#### 2.1. Transmission Anxiety

Although the stigma associated with HIV was linked to transmission, it had broader impacts on how PLHIV perceived themselves and shaped fears for their future. Participants voiced concerns such as “I don’t know what will happen to me” “what the future will bring”. Much of this sentiment derived from the way that HIV shaped people’s experiences with current and future relationships, the topic to which we now turn.

Transmission-related HIV stigma had a profound impact on PLHIV’s expectations and experiences of sexual relationships. Participants feared transmitting HIV to their sexual partners, felt responsible for avoiding transmission, and feared being stigmatised by others for being a risk of transmitting HIV to other people. Both men and women feared transmitting HIV to others and that fear weighed on them:

> *“The only thing that scares me is the possibility of transmitting this virus*.*”-* ***Female, FGD, civil society group***
>
> *“I can’t sleep with someone without a condom” “I don’t want to infect another person with this thing” “I am scared [of infecting someone]”;-* ***Male, 40+yrs, key informant interview***

Some participants avoided relationships altogether, others chose future partners based on their HIV status, and others insisted on condom use although this was not always consistent. Some participants avoided relationships, delayed sex, or chose HIV-positive partners to manage anxiety around transmitting HIV to others.

> *“But for a couple of months, I think I had this anxiety; I believe I still have it even now because I would meet someone and then as the relationship progresses, I would always run away from the sex talk, whether he is HIV positive or negative. I’ve tried dating someone who’s HIV positive, I met him in one of those support groups you get on Facebook, and with him also I was still not comfortable, … I used to fear that we would be busy and then he will take it out also, and I don’t know what he has; and you know people worry about HIV only and I’m thinking “I can’t have two or three STD’s”, so yeah, it’s something I’m still working on, the anxiety! It’s still there*.*”-* ***female, 26-30yrs, key informant interview***

Dating other PLHIV was one way participants sought to manage transmission anxiety:

> *“Well, I think I still fear infecting someone else because I still use HIV as a criteria, like a searching criteria so when I look at my list I’m like “Okay God at least he must be HIV positive also”, I have blocked the idea of dating someone who’s HIV negative in my head because I felt that it’s going to be uncomfortable for them whenever I have to take the pill in from of them; if I tell him that “this week I’m going through pill fatigue” they wouldn’t understand, instead they would think “You are being selfish, take your pill, you must protect me also”*. ***Female, 26-30yrs, key informant interview***

One female PLHIV even rejected an HIV-negative partner because she didn’t feel comfortable being intimate with him and didn’t want to “waste his time”.

> *“I remember this one guy who came to my workplace, we met, we had a conversation and he asked me out. Two weeks later, because I felt “I’m falling for this person”, I used my [HIV] status to now push him away, and he just said, “So what?” But we broke up because I wasn’t ready to get intimate, I just told him “I don’t want to waste your time, I don’t think I’ll be able to be intimate with you anytime soon so I’d rather set you free now”; at first, he could understand, I said no. let me deal with myself first and then I will try again but for now NO”* ***Female, 26-30yrs, key informant interview***

Some participants used condoms to avoid transmission:

> *“Yes it (relationship) changed. I now realised that I always have to use a condom whenever I engage in sex*.*”-* ***Female, 18-25yrs, key informant interview***
>
> *“He does understand but sometimes it becomes a challenge because his work requires him to travel and I am not sure if he complies with his treatment therefore I prefer for us to always use protection*.*”-* ***female, 18-25yrs, key informant interview***

However, condom use was inconsistent, as some participants had partners who did not want to use condoms.

> “*He refused to use a condom first of all. Obviously that led to me falling pregnant. Before falling pregnant, I experienced a lot of STI’s. One of them being HPV, Syphilis, Gonorrhoea, I had a whole list of them, that’s what took me to the clinic at the end of the day, at the beginning, before I even fell pregnant. So, I was in and out of the clinic, my parents didn’t even know that I went to the clinic” –****– female, FGD, civil society group***

Others recognized that condoms could not be used if partners were trying to conceive.

*“When it comes to my future partner, we will use protection, but when the time comes then he decides we should have a baby, I would then ask him to come with me to the clinic. He will receive counselling, and maybe they will tell him how and what to do*.*”* ***Female, 26-30yrs, key informant interview***

#### 2.2. Reputation, fear of rejection, and challenges finding a partner

Participants expected judgment and rejection from potential partners, friends and their families, and worried that peers would gossip about their status. These fear and negative experiences added to disclosure challenges. Some participants chose not to disclose their status manage fear of rejection, while others saw early disclosure as imperative to identifying and protecting a supportive partner. Disclosure fears also affected ART adherence, as some people felt they had to hide their medication or not take their medication if they were going out with friends. Some PLHIV feared that their HIV status would be weaponized against them via gossip or perceived shaming.

> *“If sometimes I fight with someone, maybe a relative. They tell you about your status [reveal my status to others], that’s why I don’t want to tell them*.*”* ***–Male, 40+yrs, key informant interview***

Participants bristled at the loss of privacy if their HIV status became known to their peers.

> *“Some were supportive though some were judgemental, like one of my friends would just ask me randomly “Hey lady do you take your treatment”? I would just respond and say “You are not going to tell me that I must take my treatment because I know my status”-* ***Female, 26-30yrs, key informant interview***

Many participants worried that their HIV status would reduce their ability to find relationship partners. These concerns were often based on direct experience including past rejections.

> *“And I was scared dating*… *partners rejected me*… *who would take me [as a partner] with my status”-****Female, 26-30yrs, key informant interview***

And fear of rejection led some PLHIV to think twice about entering a relationship.

> *“Right now there is this guy who is showing interest in me. I was still thinking about, if I agree to start a relationship with him, will he understand my situation? I will have to explain my situation to him. I don’t know if he will accept it or will just carry on with his life” –* ***young female FGD, civil society group***

Pessimism about finding a supportive partner was associated with a negative future outlook. Some PLHIV saw key life milestones – having a long-term relationship with a desirable partner, having children – as out of reach because of their status.

> *“I don’t want to lie, I am scared because I’m still very young and I don’t know what life has for me, and what kind of partner am I going to find, is he going to accept it or not and also I don’t have a child so I don’t know, I don’t want to lie I really do not know. However, that will never stop me from taking my treatment*.*” –****Female, 26-30yrs, key informant interview***
>
> *“At the time I thought I wouldn’t be able to have kids” -* ***Female, 26-30yrs, key informant interview***

#### 2.3. Disclosure challenges

Disclosing one’s HIV status can be a difficult conversation, and some participants – particularly younger participants sought to avoid it. For example, one female participant described “disclosing”, using an indirect method, by taking her medication in front of her partner, without actually talking about her HIV status or how she and her partner would navigate it:

> *“[Our relationship] has changed because we didn’t talk for quite some time. He stays in Durban and at the time I was in the Eastern Cape. We didn’t talk and he didn’t want me to visit him. He then decided around December that I can visit him. We never spoke about it, I used to take my treatment in front of him, I don’t know if he would hide his or what but I would take mine at the time when I had to. We are okay now, we communicate*.*” -* ***Female, 18-25yrs, key informant interview***

A young male participant describes a similar scenario in which disclosure occurred indirectly rather than through direct communication. Due to its stigma, HIV remains difficult to talk about within a relationship:

> *“. In my past relationship, it happened that my girlfriend was aware that this person is going through this, she saw my tablets but didn’t have the guts to ask me. Her friend found a way to ask me and I openly told her friend. After that I realised she was distant*..*” –****Male, 18-25yrs, key informant interview***

Older participants, who were likely more relationship-experienced, saw early disclosure as a critical “screening tool” to select a partner that would support them regardless of their HIV status:

> *“On the first day I meet her I tell her I am HIV positive; it will be up to her whether she continues, or she leaves*.*” –****Male, 40+yrs, key informant interview***

### 3. Understanding of U=U/TasP and its role in self-image and relationships

Our study respondents were recruited from two populations: PLHIV referred from public sector clinics and PLHIV identified through a civil society organization that works on TasP/U=U. Most participants recruited from the clinic were not aware, or were not confident, that ART leading to viral suppression prevents HIV transmission. For example one participant recruited from the clinic reported:

> *“Yes I’ve heard that on Facebook but they said after some time when taking your treatment you can’t transmit. But, I don’t know how true that is*.*”-* ***Female, 26-30yrs, key informant interview***

PLHIV who knew about U=U/TasP and had confidence in the science described the feeling of knowing that they would not infect their partner so long as they stayed on their medication. The quotes convey a significant reduction in anxiety, the sense of a weight being lifted, and the way that U=U/TasP opened up opportunities to live a normal life.

> *“If you stick to your treatment you don’t have to worry about infecting your partner. [Being virally suppressed,] I feel normal. I feel like anyone else. I can start a relationship with anyone I want*.*”* ***Female, 26-30yrs, civil society group***
>
> *“I was so much looking forward to the day when I would find out I was virally suppressed; my goal was to see the VL going down; when it was <40 copies, I knew I couldn’t infect someone else. Now I didn’t have to worry about HIV” “you see a very bright future”. [respondent was privy to the science of U=U early because he was in HIV research];” -* ***Male, FGD, civil society group***
>
> *“I’m not worried about transmitting to someone else because I know that when I’m virally suppressed I can’t transmit, and I AM virally suppressed*.*” –****Male***, ***civil society group***

Having information on TasP/U=U also armed PLHIV with the confidence to have more open conversations about HIV within their relationship.

> *“I disclosed my status to my baby daddy, and then*… *he was scared and it was heavy, ‘cause you know, men are weak. I’m sorry to say that. But men are weak. To understand that they have fear over a lot of things. He was very much concerned about the child again. But I got information, I educated him about U=U, and how we are going to save the child from getting infected by the virus. I was taking my treatment, I was using the condom when having sex. That was the only thing that saved my child from getting HIV” –****Female, 18-25, key informant interview***

Finally, the ability to control transmission risk through daily ART adherence led to a reduction in internalized stigma and the sense that PLHIV could be fully moral actors in the world.

*“being virally suppressed means I’m a full human again; HIV is no longer an issue; what is for me is to keep it suppressed;*…*I didn’t feel comfortable shaking hands initially, but now* …*I can really stand up and just be myself; I can talk about HIV without feeling like I’m a victim”*.*-* ***Male, FGD, civil society group***

## DISCUSSION

This paper aimed to look at how do PLHIV experience their HIV status in the UTT era and whether the meaning of HIV evolved as the epidemic matured. We conducted interviews and focus groups with PLHIV in Johannesburg, South Africa, in May 2021 to better understand contemporary experiences of people living with the virus.

While many participants view HIV as “normal”, due to the availability of life-saving ART and increased visibility of PLHIV, transmission-related stigma still remains pervasive. Our findings echo those of other studies that have found that stigma remains prevalent despite the scale up of treatment [31, 32]. This is particularly concerning as studies have shown that stigma negatively impacts testing for HIV, a barrier to ART adherence and is associated with depression among PLHIV [33, 34]. Consequently, these problems ultimately undermine efforts towards the elimination of HIV [35] and the stigma associated with it.

HIV stigma is often internalized, leading to continued psychosocial harm [6]. Our findings show that study respondents reported feelings of shame and guilt for contracting the virus and perceived their infection as a moral transgression. These findings are consistent with prior research in sub-Saharan Africa, which showed that HIV infection was perceived to be a result of risky sexual behaviour [2, 7].

Alongside the shame of contracting HIV, respondents felt a deep responsibility to avoid transmitting the virus to others. Respondents reported fear and anxiety around transmission as a major concern. Prior literature suggests that PLHIV are deeply concerned about transmission and engage in a variety of behaviours to protect partners from infection, a phenomenon known as “HIV prevention altruism” [36]. To manage transmission anxiety, participants avoided relationships, avoided sex, choose to date other PLHIV, or insisted on condom use. These results are consistent with previous findings whereby individuals reported diminished sexual desire due to fears of transmitting HIV to others [37, 38]. Engaging in preventive behaviors to limit transmission represented an important zone of “moral agency” for PLHIV. For those who knew about TasP/U=U, the ability to effectively prevent transmission through daily ART adherence was tremendously freeing and an important burden lifted.

Another major concern was fear of rejection, which manifested in disclosure challenges. Some participants chose to conceal their status due to fear of judgement and stigma from their family, friends and/or partners. In some instances, indirect disclosure of HIV status occurred despite participants efforts to conceal status. Non-disclosure due to stigma can result in additional challenges for PLHIV including interruptions in treatment and can negatively impact quality of life [39]. Conversely, several studies show that individuals who disclose their status are more likely to receive support from their loved ones and are more likely to adhere to treatment [40]. Fear of rejection was also linked to internalized stigma, with several participants conveying that partners would be well within their rights to reject them because they were HIV-positive, implying that they perceived themselves to be a less worthy partner than someone who was HIV-negative.

Most participants in our study did not have knowledge of TasP/U=U and this is consistent with recent empirical work from South Africa [25, 27] and a global systematic review [28]. In our study, the only participants that were knowledgeable about TasP were those recruited from a civil society organization that does advocacy work on TasP/U=U. These participants expressed that they were no longer worried about infecting their partners. Moreover, they felt confident disclosing their status to others because they knew that they could not infect someone else. And they no longer feared rejection because their partners had no good reason to reject them.

Our findings underscore the significant extent to which messaging on TasP/U=U may address the significant hardships experienced by PLHIV due to persistent transmission-related stigma, anxiety and shame. It is therefore paramount to ensure that U=U/TasP information is disseminated widely [41]as it offers an opportunity to reduce stigma and discrimination against PLHIV [42]. The message of U=U/TasP offers people the opportunity to exercise their moral agency, to have full lives with families, sex, and relationships without feeling like they are a threat to others. Equipping PLHIV and the society at large with sufficient knowledge is a powerful tool to eliminate misinformation and break the cycle of stigma.

The results should be considered in light of several study limitations. First, the study population is not representative of the general population. Our study was conducted in a predominantly peri-urban urban setting with only a few health facilities included. The study is subject to potential bias due to some of the participants recruited via referrals. In particular, the participants who were knowledgeable about TasP/U=U were identified through a U=U advocacy organization and may have had greater confidence in TasP than other persons knowledgeable of the science. We also did not explicitly recruit groups of people that are at higher risk for HIV acquisition including members of the LGBTQIA+ population, sex workers, and people who inject drugs. Despite these limitations, we were able recruit a study population via purposive sampling that was diverse in terms of gender, age and level of U=U/TasP knowledge. Second, although we reached saturation in our key themes, the sample size was limited, and this study may need to be replicated in other settings and augmented by quantitative surveys to further establish generalizability. Third, coding/analysis of qualitative data is subjective. To mitigate the impact of potential rater bias, we had at least two coders (TS, RC, JB) code all transcripts and held detailed discussions to reconcile differences in codes and interpretation.

## CONCLUSION

Persistent stigma in the UTT era is rooted in the sexual transmission of HIV. PLHIV experience self-stigma related to their status, experience anxiety about transmitting HIV to others, and fear rejection from partners based on their HIV status. Disseminating information on TasP/U=U could reduce the psychosocial burdens of HIV including self/internalized stigma, encourage disclosure, and remove barriers to HIV testing and treatment adherence.

## Data Availability

The authors declare that the data will be made available.

## ACKNOWLEDGEMENTS

The authors would like to thank the members of the HIV Survivors and Partners Network, and the Prevention Access Campaign.

## REFERENCES

1. Sontag S. AIDS and its metaphors. The disability studies reader. 1997:232–40.

2. Campbell C, Nair Y, Maimane S, Nicholson J. Dying Twice’ A Multi-level Model of the Roots of AIDS Stigma in Two South African Communities. Journal of health psychology. 2007;12(3):403–16.

3. Duffy L. Suffering, shame, and silence: The stigma of HIV/AIDS. Journal of the Association of Nurses in AIDS Care. 2005;16(1):13–20.

4. Hartwig KA, Kissioki S, Hartwig CD. Church leaders confront HIV/AIDS and stigma: a case study from Tanzania. Journal of Community & Applied Social Psychology. 2006;16(6):492–7.

5. Otolok-Tanga E, Atuyambe L, Murphy CK, Ringheim KE, Woldehanna S. Examining the actions of faith-based organizations and their influence on HIV/AIDS-related stigma: a case study of Uganda. African health sciences. 2007;7(1):55–60.

6. Simbayi LC, Kalichman S, Strebel A, Cloete A, Henda N, Mqeketo A. Internalized stigma, discrimination, and depression among men and women living with HIV/AIDS in Cape Town, South Africa. Social Science & Medicine. 2007;64(9):1823–31.

7. Wood K, Lambert H. Coded talk, scripted omissions: The micropolitics of AIDS talk in an affected community in South Africa. Medical Anthropology Quarterly. 2008;22(3):213–33.

8. Bos AER, Onya H. Fear of stigmatization as barrier to voluntary HIV counselling and testing in South Africa. East Afr J Public Health. 2008;5(2):49.

9. Daftary A, Padayatchi N, Padilla M. HIV testing and disclosure: a qualitative analysis of TB patients in South Africa. AIDS care. 2007;19(4):572–7.

10. Grant E, Logie D, Masura M, Gorman D, Murray SA. Factors facilitating and challenging access and adherence to antiretroviral therapy in a township in the Zambian Copperbelt: a qualitative study. AIDS care. 2008;20(10):1155–60.

11. Izugbara CO, Undie C-C, Mudege NN, Ezeh AC. Male youth and Voluntary Counseling and HIV-Testing: the case of Malawi and Uganda. Sex Education. 2009;9(3):243–59.

12. Katz IT, Ryu AE, Onuegbu AG, Psaros C, Weiser SD, Bangsberg DR, et al. Impact of HIVLJrelated stigma on treatment adherence: systematic review and metaLJsynthesis. Journal of the International AIDS Society. 2013;16:18640.

13. Mabunda G. Voluntary HIV counseling and testing: knowledge and practices in a rural South African village. Journal of Transcultural Nursing. 2006;17(1):23–9.

14. Weiser SD, Heisler M, Leiter K, Percy-de Korte F, Tlou S, DeMonner S, et al. Routine HIV testing in Botswana: a population-based study on attitudes, practices, and human rights concerns. PLoS medicine. 2006;3(7):e261.

15. Johnson LF, Mossong J, Dorrington RE, Schomaker M, Hoffmann CJ, Keiser O, et al. Life expectancies of South African adults starting antiretroviral treatment: collaborative analysis of cohort studies. PLoS medicine. 2013;10(4):e1001418.

16. Bor J, Tanser F, Newell M-L, Bärnighausen T. In a study of a population cohort in South Africa, HIV patients on antiretrovirals had nearly full recovery of employment. Health affairs. 2012;31(7):1459–69.

17. Camlin CS, Charlebois ED, Getahun M, Akatukwasa C, Atwine F, Itiakorit H, et al. Pathways for reduction of HIVLJrelated stigma: a model derived from longitudinal qualitative research in Kenya and Uganda. Journal of the International AIDS Society. 2020;23(12):e25647.

18. UNAIDS. Country factsheets South Africa 2020 [Available from: https://www.unaids.org/en/regionscountries/countries/southafrica.

19. Forum South African Private Practitioners Forum. Quality of HIV care higher in clinics than hospitals 2021 [Available from: https://sappf.co.za/news/413964.

20. NDoH. National Policy on HIV Pre-exposure Prophylaxis (PrEP) and Test and Treat (T&T). Pretoria: National Department of Health. 2016.

21. NDoH. Fast tracking implementation of the 90-90-90 strategy for HIV, through implementation of the test and treat (TT) policy and same-day anti-retroviral therapy (ART) initiation for positive patients. Pretoria, South Africa: National Department of Health (NDoH) 2017.

22. Chan BT, Tsai AC. HIV stigma trends in the general population during antiretroviral treatment expansion: analysis of 31 countries in sub-Saharan Africa, 2003–2013. JAIDS Journal of Acquired Immune Deficiency Syndromes. 2016;72(5):558–64.

23. Horter S, Bernays S, Thabede Z, Dlamini V, Kerschberger B, Pasipamire M, et al. “I don’t want them to know”: how stigma creates dilemmas for engagement with treat-all HIV care for people living with HIV in Eswatini. African Journal of AIDS Research. 2019;18(1):27–37.

24. Kalichman SC, Mathews C, Banas E, Kalichman MO. Treatment adherence in HIV stigmatized environments in South Africa: stigma avoidance and medication management. International journal of STD & AIDS. 2019;30(4):362–70.

25. Bor J, Bärnighausen T, Tanser F, Barofsky J, Flanagan D. Life plans of young adults in rural KwaZulu-Natal: intervention study. Forthcoming. 2018.

26. Mabuto T, Mshweshwe-Pakela N, Ntombela N, Hlongwane M, Wong V, Charalambous S, et al. Is HIV Post-test Counselling Aligned with Universal Test and Treat Goals? A Qualitative Analysis of Counselling Session Content and Delivery in South Africa. AIDS and Behavior. 2021;25(5):1583–96.

27. Bor J, Musakwa N, Onoya D, Evans D. Perceived efficacy of HIV treatment-as-prevention among university students in Johannesburg, South Africa. Sexually transmitted infections. 2021;97(8):596–600.

28. Bor J, Fischer C, Modi M, Richman B, Kinker C, King R, et al. Changing knowledge and attitudes towards HIV treatment-as-prevention and “Undetectable= Untransmittable”: A systematic review. AIDS and Behavior. 2021;25(12):4209–24.

29. RePORT N. Integrating U=U into HIV counseling in South Africa (INTUIT-SA) 2020 [Available from: https://reporter.nih.gov/search/Ek4tFJQaLE6aKgl6hX1l_g/project-details/10082738.

30. RePORT N. Integrating U=U into HIV counseling in South Africa (INTUIT-SA) 2021 [Available from: https://reporter.nih.gov/search/Ek4tFJQaLE6aKgl6hX1l_g/project-details/10227801

31. Mojola SA, Angotti N, Denardo D, Schatz E, Xavier Gómez Olivé F. The end of AIDS? HIV and the new landscape of illness in rural South Africa. Global Public Health. 2022;17(1):13–25.

32. Viljoen L, Bond VA, Reynolds LJ, Mubekapi□Musadaidzwa C, Baloyi D, Ndubani R, et al. Universal HIV testing and treatment and HIV stigma reduction: a comparative thematic analysis of qualitative data from the HPTN 071 (PopART) trial in South Africa and Zambia. Sociology of Health & Illness. 2021;43(1):167–85.

33. MacLean JR, Wetherall K. The Association between HIV-Stigma and Depressive Symptoms among People Living with HIV/AIDS: A Systematic Review of Studies Conducted in South Africa. Journal of Affective Disorders. 2021;287:125–37.

34. Mall S, Middelkoop K, Mark D, Wood R, Bekker L-G. Changing patterns in HIV/AIDS stigma and uptake of voluntary counselling and testing services: the results of two consecutive community surveys conducted in the Western Cape, South Africa. AIDS care. 2013;25(2):194–201.

35. Assembly UG. Political declaration on HIV and AIDS: on the fast-track to accelerate the fight against HIV and to end the AIDS epidemic by 2030. New York: United Nations. 2016.

36. King R, Katuntu D, Lifshay J, Packel L, Batamwita R, Nakayiwa S, et al. Processes and outcomes of HIV serostatus disclosure to sexual partners among people living with HIV in Uganda. AIDS and Behavior. 2008;12(2):232–43.

37. Siegel K, Schrimshaw EW, Lekas H-M. Diminished sexual activity, interest, and feelings of attractiveness among HIV-infected women in two eras of the AIDS epidemic. Arch Sex Behav. 2006;35(4):437–49.

38. Wamoyi J, Mbonye M, Seeley J, Birungi J, Jaffar S. Changes in sexual desires and behaviours of people living with HIV after initiation of ART: implications for HIV prevention and health promotion. BMC Public Health. 2011;11:633.

39. Simbayi LC, Kalichman SC, Strebel A, Cloete A, Henda N, Mqeketo A. Disclosure of HIV status to sex partners and sexual risk behaviours among HIV-positive men and women, Cape Town, South Africa. Sexually transmitted infections. 2007;83(1):29–34.

40. Atuyambe LM, Ssegujja E, Ssali S, Tumwine C, Nekesa N, Nannungi A, et al. HIV/AIDS status disclosure increases support, behavioural change and, HIV prevention in the long term: a case for an Urban Clinic, Kampala, Uganda. BMC Health Services Research. 2014;14(1):1–11.

41. Bor J, Onoya D, Richman B, Mayer KH. A Failure to Disseminate Transformative Science-HIV Treatment as Prevention, 10 Years On. The New England journal of medicine. 2021;385(25):2305–7.

42. Coyne R, Noone C. Investigating the effect of undetectable□=□untransmittable message frames on HIV stigma: an online experiment. AIDS Care. 2022;34(1):55

